# Prognostic impact of age and MDS-associated mutations in *NPM1*-mutated AML

**DOI:** 10.1101/2025.11.03.25339099

**Authors:** Vivian M. Liu, Megan Othus, Jasmine Naru, Rhonda E. Ries, Era L. Pogosova-Agadjanyan, Frederick R. Appelbaum, Thomas R. Chauncey, Eliana Dietrich, Harry P. Erba, John E. Godwin, Matthew P. Fitzgibbon, Min Fang, Stanley C. Lee, Anna Moseley, Mary-Elizabeth Percival, Guangrong Qin, Jerald P. Radich, Suravi Raychaudhuri, Cheryl L. Willman, Soheil Meshinchi, Derek L. Stirewalt

**Author notes:** Corresponding Author: Vivian Liu, MD, Fred Hutchinson Cancer Center, Translational Sciences and Therapeutics Division, 1100 Fairview Ave N., Seattle, WA 98109.

## Abstract

Nucleophosmin-1 (*NPM1*) mutations define a major molecular subtype of acute myeloid leukemia (AML) and is generally associated with favorable prognosis. However, the impact of myelodysplasia-associated mutations (MDSm+) on patient outcomes within this subgroup remains uncertain. We retrospectively analyzed 271 *NPM1*-mutated AML patients from three independent cohorts (SWOG, Fred Hutch, and Beat AML) to assess the prognostic significance of MDSm+ and its interaction with age. MDSm+ occurred in 17% of cases, most commonly involving *SRSF2* and *SF3B1*. Although MDSm+ was associated with inferior overall survival compared to MDSm-in ELN2022 favorable-risk patients (HR 2.0, *p*=0.008), this effect was largely driven by worse outcomes in older patients (_≥_65 years) as older ELN22 favorable-risk patients had poor OS regardless of presence of MDSm+ compared to younger patients. After stratification of patients by age, there was not a significant difference between MDSm+ and MDSm-in either younger patients (HR 0.99, p=0.98) or older patients (HR 1.42, *p*=0.33). These findings indicate that MDSm+ in *NPM1*+ AML is not independently associated with adverse risk after adjusting for age and highlight the need for age-adjusted AML risk models.

---

Nucleophosmin-1 mutations (*NPM1*+) are one of the most common genomic alterations in adult myeloid leukemia (AML) and are generally associated with chemosensitive disease and favorable outcomes, especially when there is not a concurrent *FLT3* internal tandem duplication (*FLT3*-ITD-). Currently, AML risk stratification is based on cytogenetic changes and genomic mutations per the European Leukemia Net model, updated most recently in 2022 (ELN2022)^1^. Recent updates to the ELN2017 and ELN2022 include the integration of genes frequently mutated in myelodysplastic syndrome, defined as mutations in *ASXL1, BCOR, EZH2, SF3B1, SRSF2, STAG2, RUNX1, U2AF1*, and *ZRSR2* (“MDS-associated” mutations; MDSm+), as an adverse risk factor. It is unclear whether MDSm+ alters the favorable-risk characteristics of *NPM1*+/*FLT3*-ITD-AML, and thus MDSm+ do not down-classify favorable-risk *NPM1*+ AML to adverse-risk disease. We have previously shown that older *NPM1*+/*FLT3*-ITD-AML patients, particularly age ≥65 years, have a poor prognosis (2-year overall survival for age 55-65 vs. >65 was 70% vs. 27%; *p*<0.001)^2^. Given that the frequency of MDSm+ increases with age, the strong correlation between age and inferior outcomes may be partially attributable to the MDSm+ in older patients with *NPM1*+ AML.

Previous studies on the impact of MDSm+ in *NPM1*+ AML patients have reported conflicting results^3-7^. Some have shown inferior outcomes in favorable risk *NPM1*+ patients with MDSm+^3,4^. In particular, Chan et al. reported that an inferior overall survival (OS) in *NPM1*+/MDSm+ disease is driven by older patients (≥60 years), although treatment was heterogeneous and included non-intensive therapies or no treatment in 24%^3^. They also showed that MDSm+ was not associated with adverse outcomes for younger patients^3^. Mrozek et al. demonstrated a significant difference in CR rate, disease-free survival, and OS in *NPM1*+/*FLT3*-ITD-patients with and without MDSm+, although they did not stratify their cohort by age^4^. In addition, a meta-analysis of 14 studies demonstrated worse OS for MDSm+ in only favorable-risk *NPM1*+ AML (though some of these studies used ELN2017 while others used the ELN2022 classification)^8^. Other studies, including a large study of over 1300 *NPM1*+ patients in the UK^5^, did not find MDSm+ were significantly associated with adverse outcomes in *NPM1*+ patients as a whole^5,7^ or in ELN2022-favorable risk *NPM1*+ patients^5,6^. Since most studies did not address the association between increasing age and prevalence of MDSm+, we evaluated the impact of MDSm+ on outcomes for adults with *NPM1*+ AML and favorable-risk disease, with particular attention to the impact of age.

We performed a retrospective analysis of 271 pre-treatment diagnostic specimens from three cohorts of NPM1+ AML patients: 1) 119 patients enrolled into SWOG clinical trials (SWOG-9031, SWOG-9333, S0106, and S0112)^9-12^ 2) 95 locally treated patients (Fred Hutch), and 3) 57 patients from the Beat AML study^13^. All patients were treated with curative-intent intensive chemotherapy regimens and had documentation of cytogenetic and long-term outcome data. Clinical outcomes and correlative studies have previously been reported for all SWOG and Beat AML patients. All studies for these analyses were performed according to the Institutional Review Board approved protocols and in accordance with the Declaration of Helsinki.

DNA was extracted from Fred Hutch and SWOG specimens with targeted genomic sequencing performed as previously described (Supplemental Methods)^14^. Genomic mutations for Beat AML were obtained from previously published data^13^. Morphologic complete remission (CR) rates were tabulated and compared between groups using logistic regression models stratified by cohort. Event-free survival (EFS) and overall survival were estimated using the Kaplan-Meier method and compared using Cox regression models stratified by cohort. Among participants who achieved a CR, cumulative incidence of time to relapse (TTR) was estimated non-parametrically and compared using cause-specific hazard models stratified by cohort.

Patient characteristics and co-occurring mutations are shown in Supplementary Table 1 and Supplementary Figure 1. MDSm+ occurred in 17% of patients, with the most common mutations being in *SRSF2* (7%) and *SF3B1* (3%) (Supplementary Figure 1). There were no significant differences between baseline characteristics of each of the three cohorts (Supplementary Table 1).

To assess the impact of MDSm+ on outcomes for *NPM1*+ AML patients, we compared clinical outcomes between patients stratified into the following groups employing ELN2022 criteria: 1) favorable risk/MDSm-(n=116), 2) favorable risk/MDSm+ (n=33), 3) intermediate risk (n=107) and 4) adverse risk (n=15). Favorable/MDSm+ cases had a significantly worse OS [HR 2.0, 95% CI (1.1, 3.3), p=0.008] compared to favorable/MDSm- and similar OS to intermediate and adverse-risk groups (**Figure 1, Table 1**). However, no significant difference was identified in EFS, time to relapse (TTR), or complete remission (CR) rates between favorable/MDSm+ and favorable/MDSm-patients (**Figure 1**, Supplementary Figure 2). These results suggest that MDSm+ may be an adverse risk factor in *NPM1*+ patients who are classified as favorable risk.

**Figure 1.**
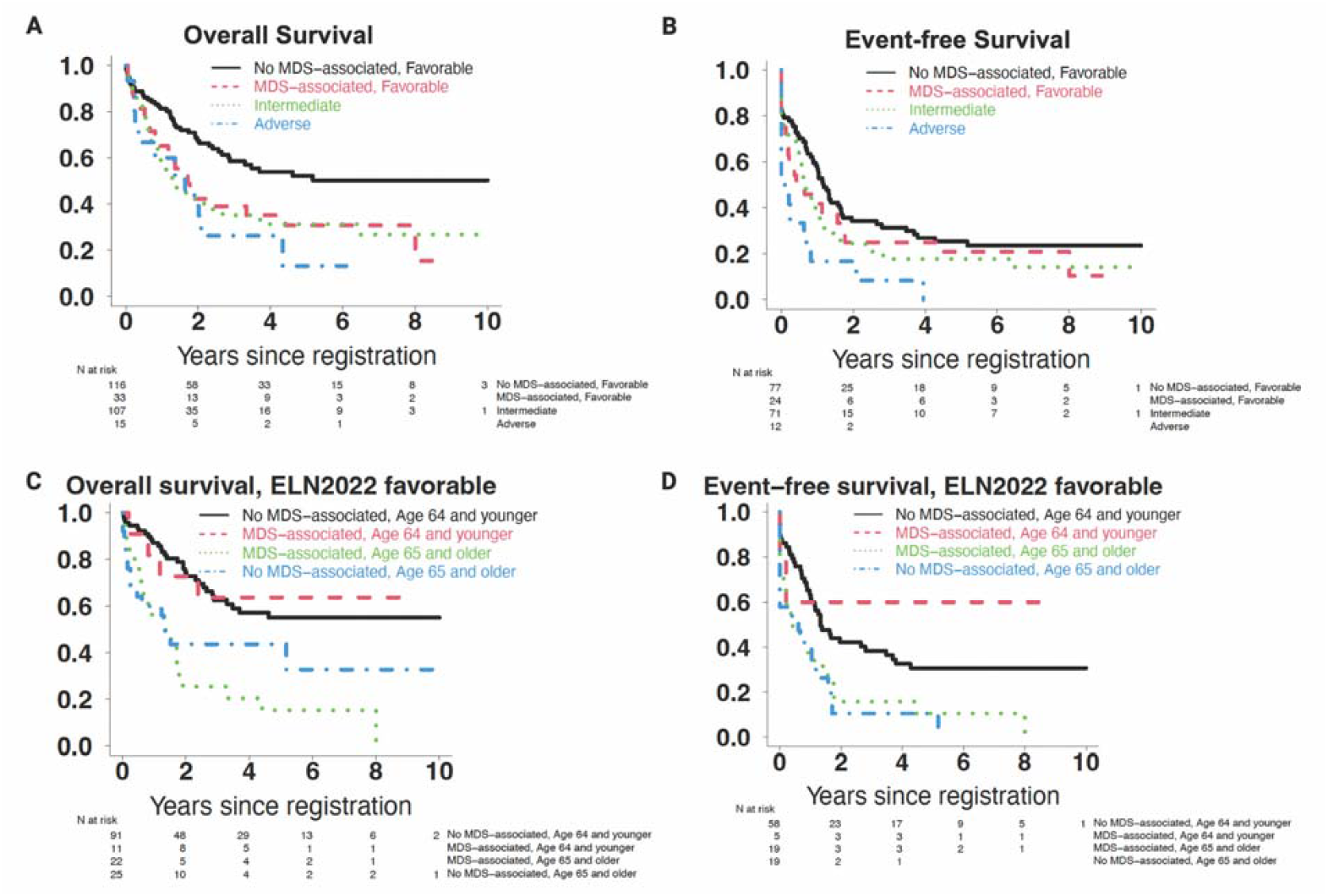
Impact of MDS-associated mutations in *NPM1*+ AML based on ELN2022 risk groups and age. Overall survival and event-free survival were analyzed for (A, B) all *NPM1*+ patients stratified by ELN2022 risk group (C, D) ELN2022 favorable-risk *NPM1*+ patients stratified by age and MDS-associated mutation.

**Table 1.** Patient outcomes in ELN2022 favorable-risk *NPM1*+ AML with and without MDS-associated mutations (MDSm+)

| **Outcome (All)** | **No MDSm** | **MDSm+** | **OR/HR** | **p-value** |
| --- | --- | --- | --- | --- |
| **n (%), N=149** | 116 (78%) | 33 (22%) | - | - |
| **CR rate (%)** | 81% | 75% | 1.35 (0.5, 3.67) | 0.56 |
| **2-year relapse (%), 95% CI)** | 57% (42-69%) | 60% (30-81%) | 1.16 (0.56, 2.39) | 0.69 |
| **2-year EFS (%), 95% CI)** | 34% (25-47%) | 25% (13-50%) | 1.27 (0.76, 2.12) | 0.37 |
| **2-year OS (%), 95% CI)** | 67% (59-77%) | 42% (28-64%) | 2.00 (1.20, 3.33) | 0.0081 |
| **Outcome (Age<65)** |  |  | **OR/HR** | **p-value** |
| **n (%), N=102** | 91 (89.2%) | 11 (10.8%) | - | - |
| **CR rate (%)** | 87% | 89% | 0.82 (0.09, 7.35) | 0.80 |
| **2-year relapse (%), 95% CI)** | 50% (34-64%) | 25% (0-71%) | 0.41 (0.05, 3.01) | 0.38 |
| **2-year EFS (%), 95% CI)** | 42% (31-57%) | 60% (29-100%) | 0.55 (0.13, 2.29) | 0.41 |
| **2-year OS (%), 95% CI)** | 74% (65-85%) | 73% (51-100%) | 0.99 (0.35, 2.83) | 0.98 |
| **Outcome (Age 65+)** |  |  | **OR/HR** | **p-value** |
| **n (%), N = 47** | 22 (46.8%) | 25 (53.2%) | - | - |
| **CR rate (%)** | 62% | 70% | 0.70 (0.19, 2.56) | 0.59 |
| **2-year relapse (%), 95% CI)** | 89% (22-99%) | 73% (32-92%) | 1.03 (0.37, 2.56) | 0.96 |
| **2-year EFS (%), 95% CI)** | 11% (3-39%) | 16% (6-45%) | 1.23 (0.63, 2.41) | 0.54 |
| **2-year OS (%), 95% CI)** | 44% (28-68%) | 25% (12-54%) | 1.42 (0.70, 2.87) | 0.33 |

MDSm+: myelodysplastic syndrome associated mutations, OR: odds ratio, HR: hazard ratio, CR: complete response, CI: confidence interval, EFS: event-free survival, OS: overall survival

Age is a potential confounder since it is both associated with the development of MDSm+ and adverse outcomes for *NPM1*+ patients^2,15^. Therefore, we further stratified *NPM1*+ patients based on age, using a cut-off threshold previously defined as being associated with adverse risk (<65 years vs. ≥65 years)^2^. Subsequent analyses showed that there was no longer a significant difference in outcomes between favorable/MDSm+ and favorable/MDSm-in both younger and older patients (**Figure 1, Table 1**). The older favorable-risk patients (≥65 years) had a significantly inferior OS and EFS compared to younger patients, regardless of presence of MDSm+, with older MDSm+ patients having the worst OS and EFS (**Figure 1, Table 1**). These results suggest that much of the previously identified adverse effect of MDSm+ could be due to the confounding variable age, and after controlling for age by risk-stratification, this effect is mitigated.

We and others have previously shown that the *NPM1*+/*FLT3*-ITD-genotype is a favorable mutational profile in patients <65 years but no longer favorable in patients ≥65 years old^2,15^. Similarly, in this study, we found that *NPM1*+ AML patients ≥65 years have adverse clinical outcomes, even in those that are ELN2022 favorable-risk without MDSm+. While *NPM1*+ AML patients harboring MDSm+ have significantly worse OS compared to MDSm-favorable-risk *NPM1*+ AML, this difference is likely driven more by age than the MDSm+.Increasing age is also highly correlated with increased prevalence of MDSm+^15^, suggesting that differences among published studies may stem from differences in cohort age, which has not been adequately addressed by prior studies. Chan et al. found significantly worse OS in *NPM1*+ AML with MDSm+ in both their full cohort and ELN2017 favorable-risk patients.

When patients were stratified by age, this effect was driven by patients who were older (defined as ≥60 years)^3^. However, their cohort included 56 patients (24.1%) who either received lower intensity therapy or no therapy, which were significantly enriched in the MDSm+ population (37.2% vs. 17.9%, p=0.008). Thus, the findings of the previous study may potentially be confounded by differences in intensity of treatment, whereas all patients in the current, larger cohort were treated with intensive induction chemotherapy. Notably, other analyses that have not found a significant difference between *NPM1*+ AML with and without MDSm+ have a lower median age in their study population. For instance, Othman et al. did not find a significant difference in *NPM1*+ AML with and without MDSm+, but the median age was 53 (range of 6 to 76 years) with only 20% of patients ≥60 years. Similarly, Ruhnke et al. had a low median age in the ELN2022 favorable group of 51 years^6^. Therefore, it is possible that the discrepancy in outcomes between various studies is driven by the difference in median age of different cohorts, with cohorts with a higher median age more likely to find a statistically significant difference.

While this study examines a large overall cohort of 271 *NPM1*+ patients, the numbers remain limited for subset analyses. For instance, there was a potential trend toward decreased OS in older favorable/MDSm+ patients, but the trend did not reach statistical significance (HR 1.42, p=0.33). We were also unable to evaluate the prognostic impact of mutations for individual MDSm+ genes given small subset numbers. In addition, data regarding allogeneic transplant and treatment after relapse were not available in many patients (e.g. SWOG), which limits our ability to assess the potential impact of subsequent therapies as a confounding variable.

Our study demonstrates that age is a strong predictor of outcomes in *NPM1*+ AML, even in ELN2022 favorable-risk disease. Older patients with ELN2022 favorable-risk *NPM1*+ AML have worse outcomes than their younger counterparts, regardless of MDSm+, suggesting that these patients should be treated more aggressively and considered for upfront allogeneic transplant if they are physically fit. The finding of MDSm+ in patients with *NPM1*+ AML is not independently associated with adverse risk after adjusting for age. Therefore, further comprehensive studies investigating the effect of age in *NPM1*+ AML are warranted to develop more precise risk-stratification guidelines.

## Data Availability

All data produced in the present study are available upon reasonable request to the authors

## Acknowledgements

This study was funded by the SWOG/Hope Foundation for Cancer Research, NIH/NCI grants RO1CA190661, R01CA160872, U10CA180888, U10CA180819, U24CA196175 andP30CA015704. V.M.L. and S.R. are supported by NHLBI award T32HL007093. We also thank the University of Washington/Fred Hutch Hematopoietic Diseases Repository for providing patient samples for locally-treated patients.

## Authorship Contributions

V.M.L., M.O., S.C.L. and D.L.S. designed the study. M.O., E.D., and G.Q. conducted statistical analysis. J.N. and E.L.P-A. were responsible for patient sample preparation and DNA extraction. R.E.R., M.P.F., S.M., and D.L.S. analyzed and provided genomic data. V.M.L. R.E.R., M.E.P.,A.M., F.R.A., T.R.C., H.P.E., J.E.G., J.P.R., S.R., C.L.W. and M.F. provided patient clinical data and/or oversaw clinical trial development. V.M.L. and D.L.S. wrote the manuscript. All authors reviewed and edited the manuscript.

