## Supplementary Materials for "Prognostic impact of age and MDS-associated mutations in *NPM1*-mutated AML"

**SUPPLEMENTAL METHODS**

***Identification of Genomic Mutations***

Paired-end reads were first aligned to the human genome reference assembly (GRCh37/hg19) using Burrows-Wheeler Aligner (BWA, v0.7.12) as previously described.(1) The resulting alignment data were processed according to the best practice of Genome Analysis Toolkit (GATK, v3.5 https://www.broadinstitute.org/gatk/). Sequence alignment statistics and coverages were computed using Samtools (version 1.0, http://samtools.sourceforge.net) and GATK DepthOfCoverage, respectively. Variants were called per sample using GATK HaplotypeCaller in GVCF mode, then jointly as a cohort using GenotypeGVCFs. The resulting collection of variants, in the form of a VCF file, were annotated using Annovar (version 2016Feb01). Sequencing depth for each read loci were calculated to determine average percent coverage. For quality control, loci with >20% of samples displaying <65 read coverage were removed from downstream analyses. The following were excluded as potential mutations: a) synonymous variants, b) alterations with low quality (Qual score <200), variant read depth <75, and variant allele frequency (VAF) <3%; and c) alterations in non-exonic loci outside of splice sites. Additional inclusion and exclusion criteria were then applied to the remaining alterations. All nonsense and those missense alterations known to be pathogenic or likely pathogenic in ClinVar were classified as mutations, while missense alterations documented as benign or likely benign were excluded. Missense changes described as having uncertain significance, conflicting interpretation, or not reported in ClinVar were further filtered using the following in silico bioinformatic tools: (a) AlphaMissense (https://alphamissense.hegelab.org/),(2) (b) VEST (https://cravat.us/CRAVAT/),(3, 4) (c) PolyPhen,(5) (d) SIFT,(6) and (e) Provean.(7) AlphaMissense scores (≥0.564) was utilized as the initial screen for potential pathogenicity due to its improved performance,(2) but to further reduce false positives, at least one additional of the Insilco bioinformatic tools also had to score the missense as likely deleterious, damaging, and/or pathogenic using the following criteria: VEST ≥ 0.5, PolyPhen ≥ 0.5, SIFT ≤ 0.05 and/or Provean ≤ -2.5. Those missense alterations not meeting these criteria were deemed variant of unknown significance (VUS) and not consider in downstream analyses. Similarly, all inframe_indels were screened for significance in ClinVar, with those indel alterations classified as pathogenic or likely pathogenic declared mutations, while alterations documented as benign or likely benign were excluded. Those remaining indels were evaluated by VEST-indel, and those with a score ≥ 0.500 declared as likely mutations while others were classified as VUS; the latter again were not included in downstream analyses. For splice site variants, we utilized the SpliceAI_Score (8) and Pangolin_Score (9) (https://spliceailookup.broadinstitute.org/). To reduce false positives, the alteration in the splicing region had to be considered as adversely impacting the potential splice using both tools: SpliceAI_Score AND Pangolin_Score ≥ 0.75, with all other alterations being declared VUS and not utilized for analyses.

**Supplementary Table 1:** Cohort characteristics summary. N (%) and median (range) reported

| Factor | BeatAML (n=57) | Fred Hutch (n=94) | SWOG (n=120) | All | *p*-value | N miss |
|---|---|---|---|---|---|---|
| Age (years) | 57 (26, 74) | 60.5 (21, 81.68) | 56.6 (19.4, 81.4) | 58.4 (19.4, 81.68) | 0.25 | 0 |
| Age category | | | | | | |
| Age 64 and younger | 42 (74) | 57 (61) | 89 (74) | 188 (69) | 0.081 | 0 |
| Age 65 and older | 15 (26) | 37 (39) | 31 (26) | 83 (31) | | |
| Sex | | | | | | |
| Male | 26 (46) | 48 (51) | 60 (50) | 134 (49) | 0.79 | 0 |
| Female | 31 (54) | 46 (49) | 60 (50) | 137 (51) | | |
| ELN2022 risk | | | | | | |
| Favorable | 31 (54) | 50 (53) | 68 (57) | 149 (55) | 0.71 | 0 |
| Intermediate | 24 (42) | 40 (43) | 43 (36) | 107 (39) | | |
| Adverse | 2 (4) | 4 (4) | 9 (8) | 15 (6) | | |
| MDS-associated mutations | | | | | | |
| MDS-associated mutation | 8 (14) | 16 (17) | 22 (18) | 46 (17) | 0.82 | 0 |
| No MDS-associated mutation | 49 (86) | 78 (83) | 98 (82) | 225 (83) | | |

### Supplementary Figures

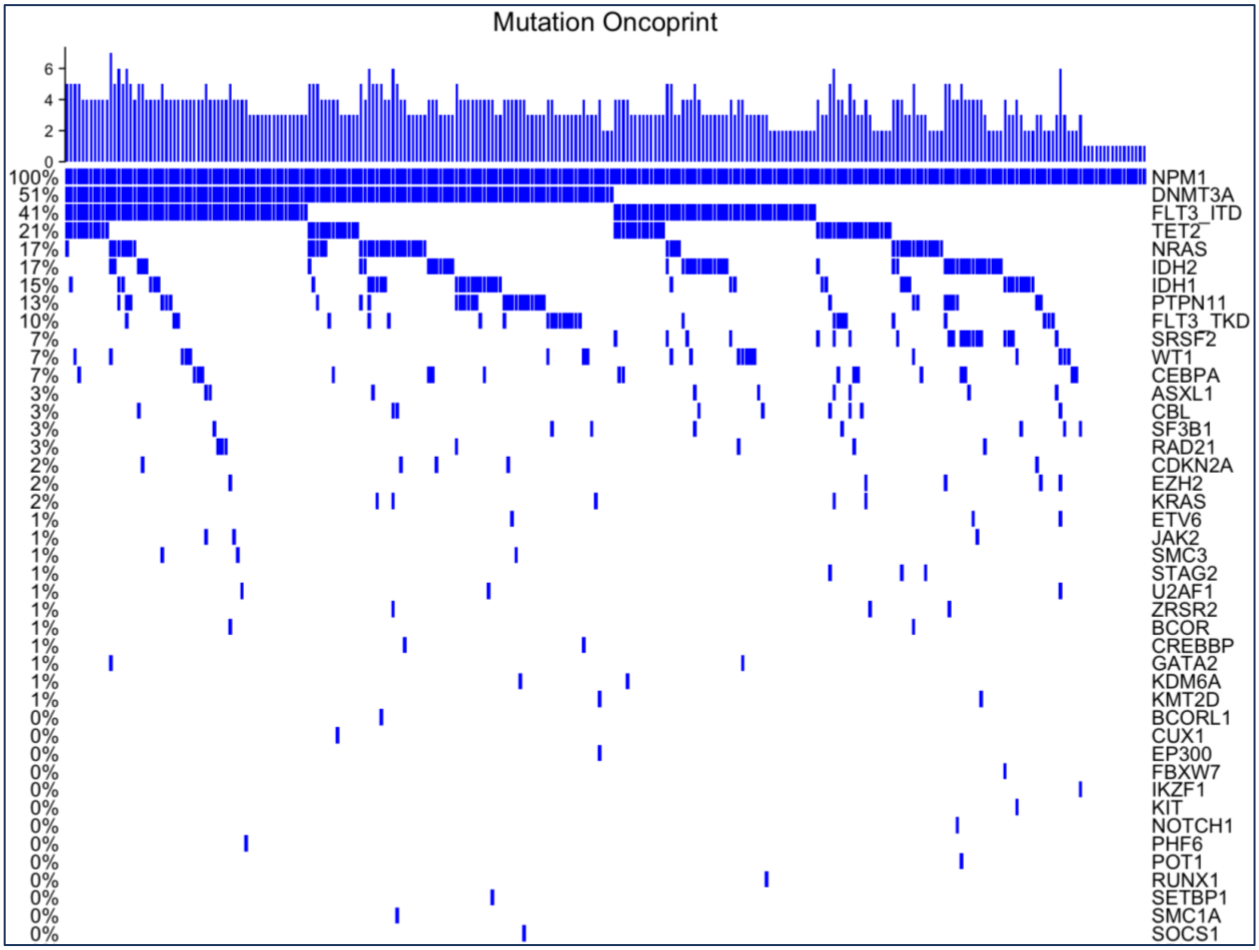


**Supplementary Figure 1:** Oncoprint of genomic mutations of entire cohort

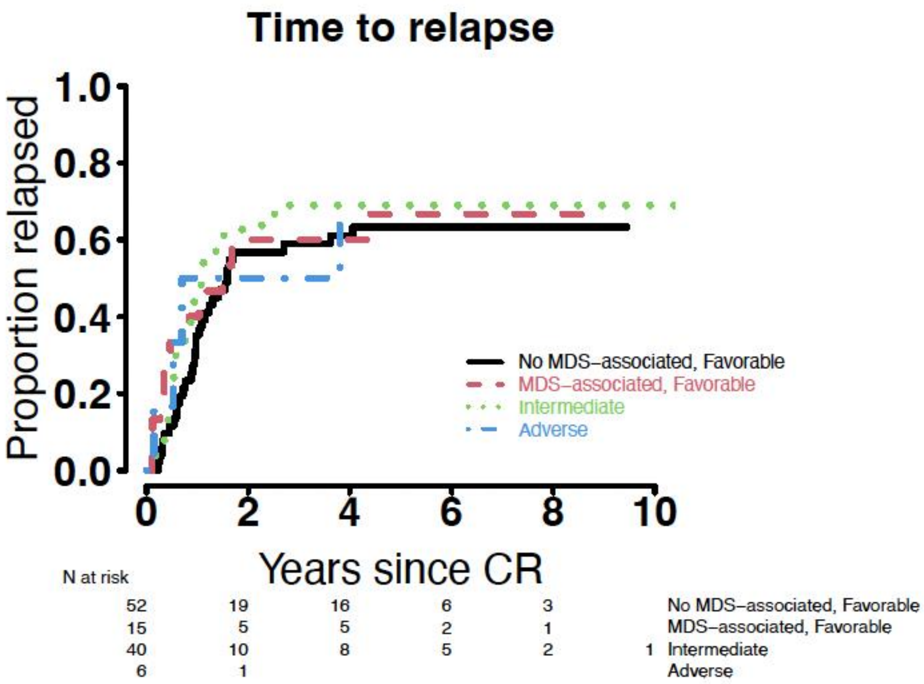


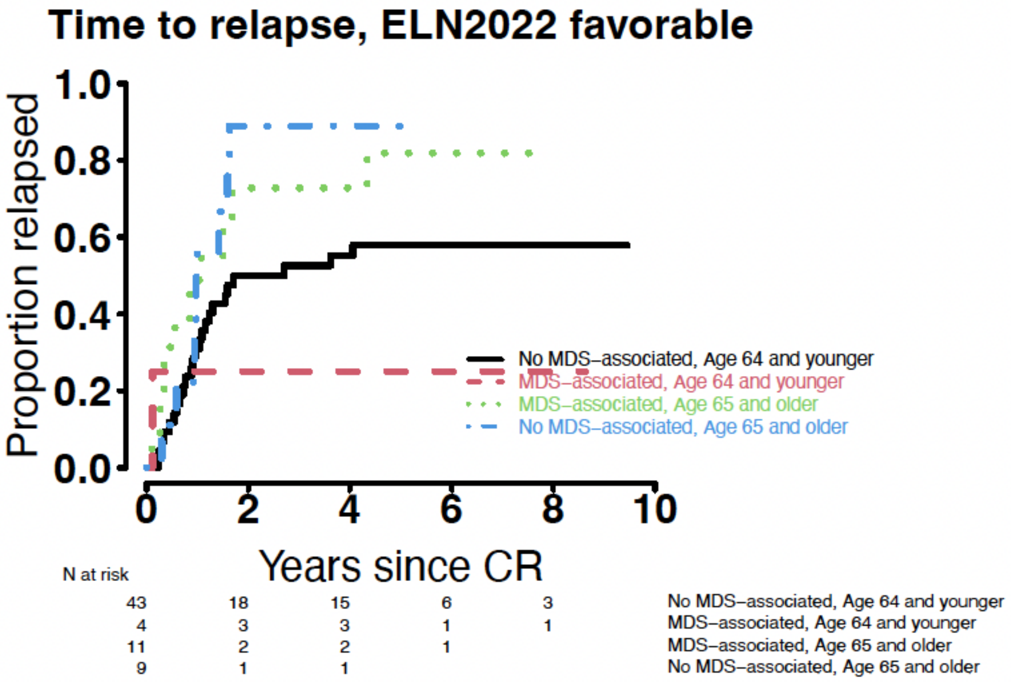


**Supplementary Figure 2:** Time to relapse for *NPM1*+ AML stratified by (A) ELN2022 risk groups (B) ELN2022 favorable-risk patients stratified by age and MDS-associated mutation.
